# Beyond predicting the number of infections: predicting who is likely to be COVID negative or positive

**DOI:** 10.1101/2020.04.30.20086348

**Authors:** Stephen X. Zhang, Shuhua Sun, Asghar Afshar Jahanshahi, Yifei Wang, Abbas Nazarian Madavani, Maryam Mokhtari Dinani

## Abstract

This study provides the first attempt to identify people at greater risk of COVID-19 infection, enabling more targeted infectious disease prevention and control, which are especially important in the ongoing shortage of COVID-19 testing.

We conducted a primary survey of 521 adults on April 1-10, 2020 in Iran, where the official infection rate was 0·08%. In our sample, 3% reported being COVID-19 positive and 15% were unsure of their status. This relatively high positive rate enabled us to conduct the analysis at the 5% significance level.

At the time of the survey, 44% of the adults worked from home; 26% still went to work in their workplaces; 27% had stopped working due to the COVID-19 pandemic; and 3% were unemployed. Adults who exercised more were more likely to be COVID-19 negative. Each additional hour of exercise per day predicted a 78% increase in the likelihood of being COVID-19 negative. Adults with chronic medical illnesses were 48% more likely to be COVID-19 negative. In terms of work situation, those who worked from home were the most likely to be COVID-19 negative, and those who had stopped working were the most likely to be COVID-19 positive. Individuals in larger organizations were less likely to be COVID-19 positive.

Given the testing shortage in many countries, we identify a novel approach to predict the likelihood of COVID-19 infection by a set of personal and work situation characteristics, in order to help to identify individuals with more or less risk of contracting the virus. We hope this research opens a new research avenue to identify the individual risk factors of COVID-19 infection to enable more targeted infectious disease prevention, communication, testing, and control to complement the effort to expand testing capacity.

## INTRODUCTION

COVID-19 (coronavirus disease 2019) is overwhelming clinical capacities in many countries. To contain the spread of COVID-19, we need to ramp up efforts to identify and isolate people with the disease early on. As the Director-General of WHO called, “We have a simple message for all countries: test, test, test”.^1^ However, even many developed countries are experiencing severe shortages of test kits.^2^ Given the limited testing capacities, it is useful to identify the groups of people who are at greater risk of contracting COVID-19 to enable more targeted infectious disease prevention, communication, testing, and control. The identification of higher risk groups can reduce the risk not only to these individuals but also to the medical system and society at large.

Unfortunately, we have limited knowledge about the predictors of who is at greater risk of COVID-19 infection. Many models have been published to predict the number of people infected by COVID-19, 3-5 but not who is more likely to be infected. Other research identified who would be more likely to develop severe symptoms once they contract the COVID infection (retrieved from US CDC).^6^ This knowledge has already prompted governments and NGOs to take preventive measures tailored to those identified groups of people, such as older people and people with chronic disease.^7,8^ New evidence has emerged that homeless people and people in care homes are at greater risk of contracting COVID-19.^8,9^ Had this been known earlier, more lives could have been saved by more targeted preventive measures.^10^

The purpose of the study is to further our knowledge of the risk factors to allow early identification of individuals more susceptible to COVID-19 infection. These predictors can help target infectious disease prevention and control towards higher risk groups. This knowledge is especially critical to countries experiencing a shortage of test kits. To explore potential predictors we conducted this study in Iran, a hotspot of COVID-19 with a shortage of test kits.^11^ We predict individuals’ likelihood of COVID-19 infection based on: (1) demographic variables, including chronic medical conditions and exercise hours;^12,13^ (2) a set of employment status and work situation variables which can affect individuals’ daily routines and movements and hence the risk of contracting the disease;^13,14^ and (3) a pair of psychiatric variables on depression and anxiety, due to their effect on how people cope with adversity.^15,16^ Such knowledge on the predictors opens a new research direction and avenue to identify individuals at greater risk of contracting COVID-19 to manage the pandemic with the shortage of test kits.

## METHODS

COVID-19 hit Iran early and hard, and Iran has been one of the countries most affected by COVID-19 since March 2020. Healthcare modeling in late March, when we designed the study, estimated the COVID-19 crisis in Iran would reach its peak in the first week of April. Accordingly, we surveyed adults in Iran on April 1-10, 2020. On April 1, official statistics reported 47,593 confirmed cases and 3,036 deaths with COVID-19. On April 10, official statistics reported 68,192 confirmed cases and 4,232 deaths with COVID-19. Overall, 0·08% of Iranians were positive for COVID-19 at April 10.

Survey participants reported their individual COVID-19 infection status as negative; unsure; or positive. Participants also reported whether they had chronic health issues (no; unsure; yes), exercise hours per day in the past week, working situation (worked from home; worked in workplace; stopped work due to COVID-19; unemployed), and Patient Health Questionnaire 4-item (PHQ-4) scale which measures depression and anxiety, as well as their demographic variables such as their gender, age, and the size of their work organization (0 for the unemployed), because large organizations were deemed to offer better healthcare coverage for their employees in Iran.

Participation was entirely voluntary and anonymous, and we distributed the survey through social media (Telegram, Instagram and WhatsApp) given the lockdown situations. The survey was approved (IR.SSRI.REC.1389.685) by the ethics committee of Shahid Rejaee University in Iran.

### Statistical Analysis

We analyzed the data using STATA 16·0 with a significance level at 0·05. Because the outcome variable of individual COVID-19 infection is ordinal (negative; unsure; positive), we predicted it by ordered logistic regressions using the STATA command of gologit2. Accordingly, the predictions of individual COVID-19 infection are in odds ratios (ORs).

## RESULTS

### Descriptive Findings

Table 1 contains the descriptive findings. Of the 521 adults who completed the survey, about half were male (49%). The average age was 43·9 years (st.d. 11·7; min: 20; max: 79). At the time of the survey, 44% of the adults worked from home; 26% still went to work in their workplaces; 27% had stopped working due to COVID-19; and 3% were unemployed. The median number of employees in a workplace was 28 (mean: 201·5; st.d. 456·8). Most participants (87%) did not have chronic medical issues; 3% were unsure whether they had chronic medical issues, and the remaining 10% had chronic medical issues. In terms of exercise hours per day in the past week, 56%, 37%, 4%, 1%, and 2% of them exercised 0, 1, 2, 3, and 4 or more hours per day, respectively. The scores on the PHQ-4 for depression and anxiety were 1·7 (st.d. 1·4) and 1·6 (st.d. 1·5) respectively, meaning 22·3% and 21·5% surpassed the cutoff levels of psychiatric screening for depression and anxiety disorders respectively. In terms of COVID-19, 82% of the participants indicated they did not have COVID-19, 15% were unsure, and 3% reported they were infected by COVID-19.

**Table 1.**
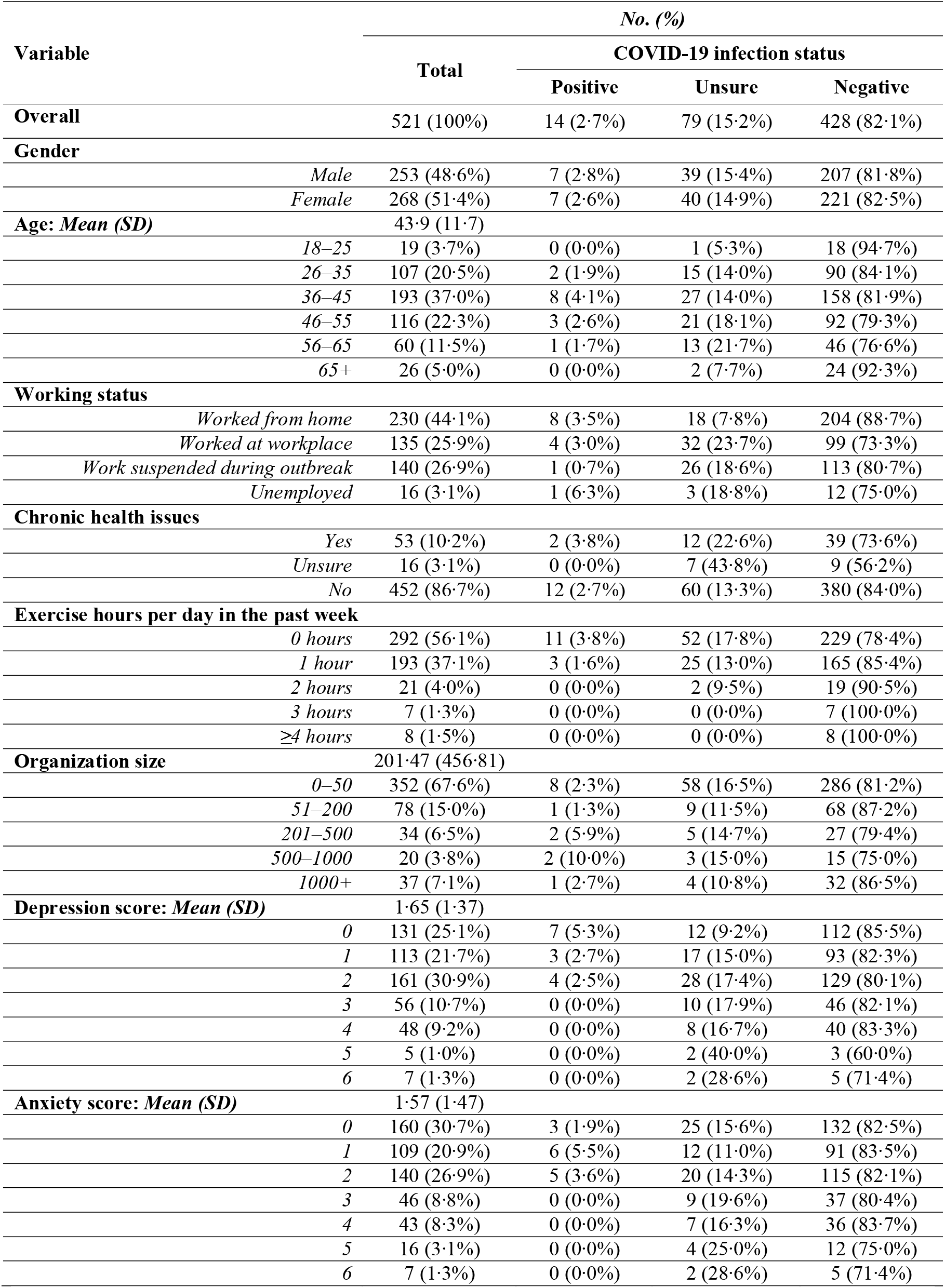
Demographic characteristics and COVID-19 status of the participants (*n*=521)

### Risk Predictors of Individual COVID-19 Infection Status

Table 2 shows the ordered logistic regressions analysis predicting the likelihood of being COVID-19 negative from the alternatives (i.e. being unsure or positive). Adults with chronic medical issues were 48% more likely to be COVID-19 negative (OR: 1·48; 95% CI: 1·06 to 2·08; p = 0·023), possibly due to them being more cautious, suggesting people had taken seriously the information on the higher fatality rate of people who had comorbidities. Adults who exercised more hours per day were more likely to be COVID-19 negative (OR: 1·78; 95% CI: 1·21 to 2·62; p = 0·003). Each additional hour of exercise per day predicted a 78% increase in the likelihood of being COVID-19 negative.

**Table 2.**
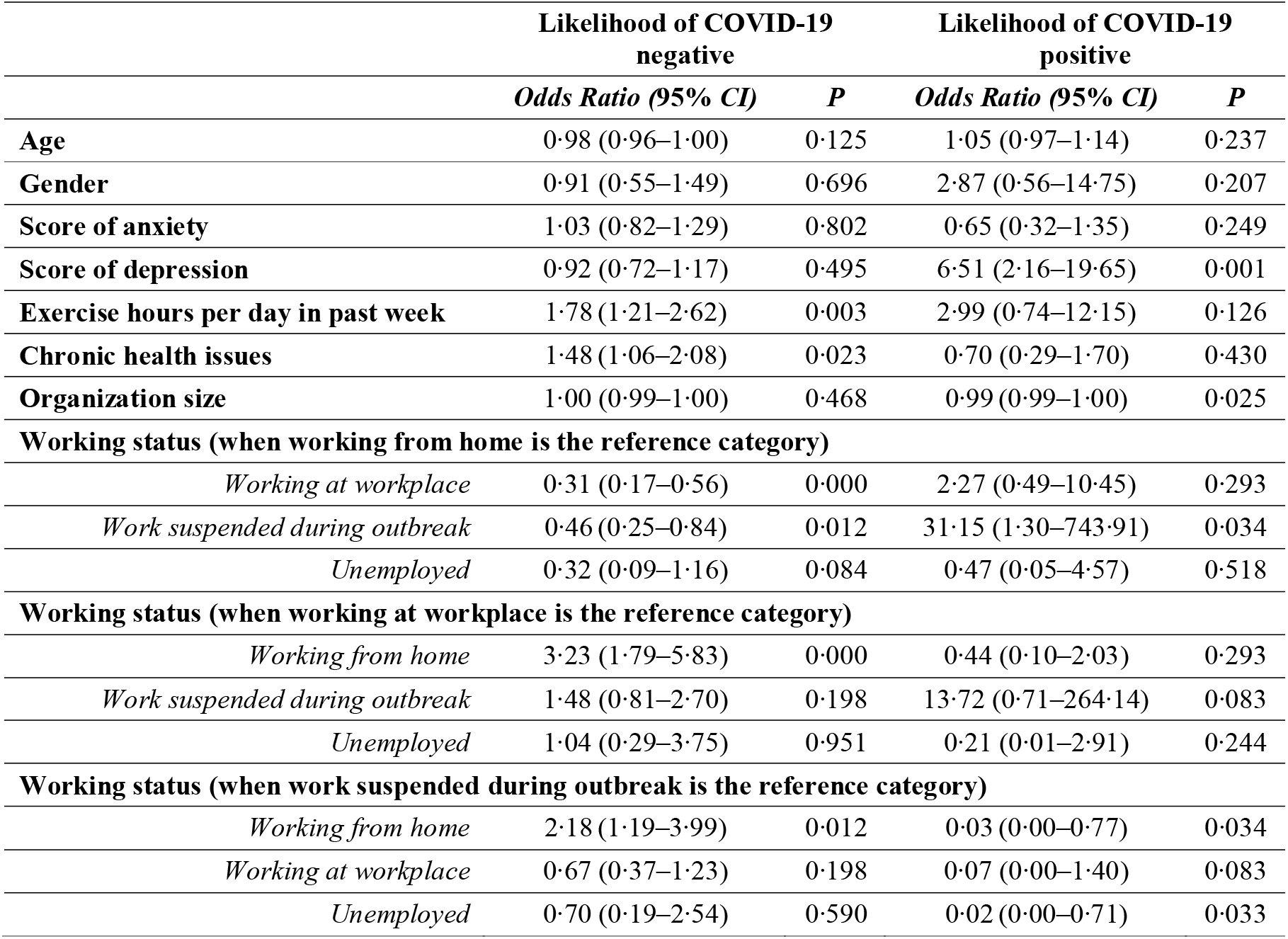
Ordered logistic regressions results predicting individuals’ likelihood of being COVID-19 positive or negative (n=521)

|  | Likelihood of COVID-19 negative |  | Likelihood of COVID-19 positive |  |
| --- | --- | --- | --- | --- |
|  | *Odds Ratio (95% CI)* | *P* | *Odds Ratio (95% CI)* | *P* |
| **Age** | 0.98 (0.96–1.00) | 0.125 | 1.05 (0.97–1.14) | 0.237 |
| **Gender** | 0.91 (0.55–1.49) | 0.696 | 2.87 (0.56–14.75) | 0.207 |
| **Score of anxiety** | 1.03 (0.82–1.29) | 0.802 | 0.65 (0.32–1.35) | 0.249 |
| **Score of depression** | 0.92 (0.72–1.17) | 0.495 | 6.51 (2.16–19.65) | 0.001 |
| **Exercise hours per day in past week** | 1.78 (1.21–2.62) | 0.003 | 2.99 (0.74–12.15) | 0.126 |
| **Chronic health issues** | 1.48 (1.06–2.08) | 0.023 | 0.70 (0.29–1.70) | 0.430 |
| **Organization size** | 1.00 (0.99–1.00) | 0.468 | 0.99 (0.99–1.00) | 0.025 |
| **Working status (when working from home is the reference category)** |  |  |  |  |
| *Working at workplace* | 0.31 (0.17–0.56) | 0.000 | 2.27 (0.49–10.45) | 0.293 |
| *Work suspended during outbreak* | 0.46 (0.25–0.84) | 0.012 | 31.15 (1.30–743.91) | 0.034 |
| *Unemployed* | 0.32 (0.09–1.16) | 0.084 | 0.47 (0.05–4.57) | 0.518 |
| **Working status (when working at workplace is the reference category)** |  |  |  |  |
| *Working from home* | 3.23 (1.79–5.83) | 0.000 | 0.44 (0.10–2.03) | 0.293 |
| *Work suspended during outbreak* | 1.48 (0.81–2.70) | 0.198 | 13.72 (0.71–264.14) | 0.083 |
| *Unemployed* | 1.04 (0.29–3.75) | 0.951 | 0.21 (0.01–2.91) | 0.244 |
| **Working status (when work suspended during outbreak is the reference category)** |  |  |  |  |
| *Working from home* | 2.18 (1.19–3.99) | 0.012 | 0.03 (0.00–0.77) | 0.034 |
| *Working at workplace* | 0.67 (0.37–1.23) | 0.198 | 0.07 (0.00–1.40) | 0.083 |
| *Unemployed* | 0.70 (0.19–2.54) | 0.590 | 0.02 (0.00–0.71) | 0.033 |

As we captured four work situations (worked from home, worked at workplace, stopped work, and unemployed), we introduced each work situation into the regression as a reference group one by one to conduct a pairwise comparison. Compared with those who worked from home, those who worked at workplace or stopped work were respectively 69% (OR: 0·31; 95% CI: 0·17 to 0·56; p = 0·000) and 54% (OR: 0·46; 95% CI: 0·25 to 0·84; p = 0·012) less likely to be COVID-19 negative. In other words, those who worked from home were more likely to be COVID-19 negative than those who went to work at workplace or had stopped working in Iran.

We further performed ordered logistic regressions analysis to predict the likelihood of being COVID-19 positive from the alternatives. As expected, depression was positively associated with the likelihood of being COVID-19 positive (OR: 6·51; 95% CI: 2·16 to 19·65; p = 0·001), but the association does not imply causality. The pairwise comparison by work situations revealed that the likelihood of being COVID-19 positive among those who had stopped working is 31·15 times those who worked from home (OR: 31·15; 95% CI: 1·30 to 743·91; p = 0·034) and 65·79 times those who were unemployed (OR: 65·79; 95% CI: 1·41 to 3069·98; p = 0·033). The p-values were significant but the confidence intervals were large due to the small number of participants who reported being COVID-19 positive. The results imply that those who had stopped work had a higher infection rate, perhaps either because they were agitated or restless now without work or had riskier jobs to begin with and had to stop working. The size of the work organization (i.e., number of employees) negatively predicted the likelihood of being COVID-19 positive (OR: 0·99; 95% CI: 0·995 to 1·000; p = 0·025), suggesting those who worked in larger organizations were safer.

It is worth noting that the predictors of being COVID-19 positive differed from the predictors of being COVID-19 negative. Moreover, variables for age, gender, and anxiety did not predict individual COVID-19 infection status at the time of the survey.

### Predicted likelihood of COVID-19 status by work situation

We also report the predicted likelihood of being COVID-19 negative, unsure, or positive by an individual’s work situation, holding the other variables constant (Figure 1). Individuals who worked from home had an 89.5% (OR: 0·895; 95% CI: 0·856 to 0·933; p = 0·000) likelihood of being COVID-19 negative, 0·6% (OR: 0·006; 95% CI: -0·069 to 0·080; p = 0·878) likelihood of being unsure, and 9·9% (OR: 0·099; 95% CI: 0·025 to 0·173; p = 0·009) likelihood of being COVID-19 positive. Overall, those who worked from home were relatively aware of their COVID-19 infection status, no matter if it was positive or negative.

Individuals who worked in the workplace had a 73·4% (OR: 0·734; 95% CI: 0·659 to 0·809; p = 0·000) likelihood of being COVID-19 negative, 20·4% (OR: 0·204; 95% CI: 0·130 to 0·279; p = 0·000) likelihood of being unsure, and 6·1% (OR: 0·061; 95% CI: 0·022 to 0·101; p = 0·002) likelihood of being COVID-19 positive. Hence, over 20% of those who went to their workplace were unsure of their COVID-19 infection status, suggesting this group of people were likely in a state of uncertainty.

Individuals who had stopped work had an 80·1% (OR: 0·801; 95% CI: 0·735 to 0·867;. p = 0·000) likelihood of being COVID-19 negative, 18·6% (OR: 0·186; 95% CI: 0·120 to 0·252;. p = 0·000) likelihood of being unsure, and 1·3% (OR: 0·013; 95% CI: -0·007 to 0·032;. p = 0·196) likelihood of being COVID-19 positive. A significant portion of these individuals were also unsure of their COVID-19 infection status.

Individuals who were unemployed had a 74·2% (OR: 0·742; 95% CI: 0·522 to 0·962;. p = 0·000) likelihood of being COVID-19 negative, 11·0% (OR: 0·110; 95% CI: -0·129 to 0·348; p = 0·367) likelihood of being unsure, and 14·9% (OR: 0·149; 95% CI: -0·039 to 0·336; p = 0·121) likelihood of being COVID-19 positive.

## DISCUSSION

COVID-19 test kits have been in short supply since the beginning of the COVID-19 outbreak and continue to be in critical shortage in many countries as the pandemic continues to develop. Given the insufficient testing capacity, we identify a novel approach to predict the likelihood of COVID-19 infection by individual risk factors, to help to identify clusters of individuals with more or less risk of contracting the virus – a critical piece of information to enable more targeted social distancing and isolation practices to contain the virus infection, especially in areas with insufficient testing.

The empirical setting of Iran had a high population-wide COVID-19 infection rate of 0·08% in early April. In our sample of over 500 adults, 3% reported being COVID-19 positive and 15% were unsure of their status. These relatively high rates enable us to conduct the analysis.

First, the results on the predictors of being COVID-19 negative reveal that two groups were more likely to be COVID-19 negative: people who exercised more and people who had chronic medical issues. While it may appear counterintuitive that those who had chronic medical issues were more likely to be COVID-19 negative, the finding is understandable, as people with chronic medical issues likely went out less and likely had taken more action to protect themselves against COVID-19 virus due to their higher chance of becoming seriously ill or dying if they do get it. The exercise finding may reflect that healthier people are more likely to be able to exercise. The finding that those who worked from home had a higher chance of being COVID-19 negative supports the shelter-in or stay-at-home orders in many parts of the world during the pandemic.

Second, the results on the predictor of being COVID-19 positive reveals, somewhat surprisingly, those who stopped working had significantly higher chance of being COVID-19 positive than those who worked from home or were unemployed. Unlike those who worked from home, or those who were unemployed who were probably more used to not working, those who have had their work stopped due to COVID-19 all of a sudden might be more agitated or restless.^17,18^ With their daily work taken away and more spare time, they might have had risk exposures elsewhere. It is also possible that those who had their work stopped may have had a riskier job to begin with and therefore had a lower chance of being COVID-19 negative due to their previous exposures at work. Nonetheless, it highlights that we should not assume those who have stopped working are safe. They remain at higher risk than the groups who worked from home or had not been employed for a long time (before and during the COVID-19 outbreak). People working in smaller organizations were at greater risk of being COVID-19 positive, suggesting epidemiological preventions could target employees of smaller organizations more.

Lastly, past reports have indicated older people and males were more likely to have COVID-19.^19,20^ Age and gender were found to be useful predictors of the mental health of adults during the COVID-19 crisis,16,19 however they failed to directly predict either COVID-19 negative or COVID-19 positive status in our analysis.

These findings identified a number of risk factors that could enable more targeted epidemiological preventions. The risk factors can help identify people to prioritize for COVID-19 testing, should testing kits become available, or in lieu, help implement more targeted social distancing and isolation measures, or conduct more specific communications on infectious disease prevention and control to high risk groups.

This study has several limitations. While the severity of the COVID-19 crisis in Iran early on presents a setting to predict COVID-19 infection status, the number of COVID-19 positive cases in our sample remained relatively small. Similarly, the number of unemployed participants in the sample was too small to enable more analysis. While we aimed to cover a broad spectrum of adults in Iran, our sample should not be taken as a representative national sample. Also, the risk factors of COVID-19 infection are likely to differ across countries given different cultural and social practices. Lastly, our model is predictive, and we do not claim causalities.

In summary, this study provides the first attempt to explore predictors of people at greater risk of COVID-19 infection during the COVID-19 pandemic. We hope this research opens a new research avenue by identifying higher risk individuals to enable more targeted infectious disease prevention, communication, testing, and control to curtail the pandemic.

## Data Availability

Upon request

## Acknowledgments

The research was self-funded as a result of research funding freeze in many institutions due to the COVID-19 pandemic.

Authors’ roles
- S. X. Z.: Conceptualization, Investigation, Methodology, Formal analysis, Visualization, Writing – Original, Writing - Review & Editing, Supervision
- S. S.: Writing - Original Draft, Writing - Review & Editing
- A. A. J.: Investigation (data collection), Resources, Conceptualization, Writing - Review & Editing
- Y. W.: Visualization, Writing - Original Draft, Writing - Review & Editing
- N. M.: Investigation (data collection)
- M. M. D. : Investigation (data collection)

